# Clinical experience with paliperidone palmitate in a specialty hospital in the Philippines: A short report

**DOI:** 10.1101/2022.01.25.22269857

**Authors:** Amadeo A. Alinea, Carl Abelardo T. Antonio, Amiel Nazer C. Bermudez, Kim L. Cochon, Maria Fatima V. Martinez, Jonathan P. Guevarra

**Affiliations:** Department of Psychiatry Veterans Memorial Medical Center, Quezon City, Philippines; Department of Health Policy and Administration, College of Public Health, University of the Philippines Manila, Manila, Philippines; Department of Applied Social Sciences, The Hong Kong Polytechnic University, Kowloon, Hong Kong; Department of Epidemiology and Biostatistics, College of Public Health, University of the Philippines Manila, Manila, Philippines; Department of Epidemiology, School of Public Health, Brown University, Providence, RI, USA; Department of Statistics, The Chinese University of Hong Kong, Sha Tin, Hong Kong; Department of Health Promotion and Education, College of Public Health, University of the Philippines Manila, Manila, Philippines

**Keywords:** cross-sectional studies, paliperidone palmitate, Philippines, real-word effectiveness, schizophrenia

## Abstract

**Objective:** To describe the clinical outcomes related to the introduction of paliperidone palmitate in a specialty hospital in the Philippines

**Design:** Cross-sectional study among patients seen at the psychiatry service of a specialty hospital catering to veterans who were initiated on paliperidone palmitate. We reviewed and abstracted baseline patient data from the medical record of eligible patients. Outcome of treatment was collected through a one-time objective assessment of the patient by a third-party psychiatrist using the Structured Clinical Interview for Symptoms of Remission (SCI-SR) tool.

**Main Result:** A total of 30 patients were recruited for the study from August 2020 and June 2021, the majority of whom were males (80%), residents of the National Capital Region (50%), and single (20%). The median duration from schizophrenia diagnosis to initiation of paliperidone treatment was 19.50 years (IQR: 16.60 – 33.50). In eight patients (22.67%), other antipsychotic drugs were discontinued following initiation of paliperidone treatment; in the remaining 22 participants (73.33%), paliperidone was taken concurrently with other antipsychotic drugs. The median duration from the initiation of paliperidone treatment to follow-up assessment was 27.20 months (IQR: 24.73 – 30.50), with all participants having at least 6 months of treatment. At follow-up assessment, all participants were classified to be in remission.

**Conclusion:** In this study among patients with schizophrenia seen in a specialty hospital in the Philippines, we found evidence that clinical outcomes with paliperidone palmitate were comparable to those given a combination of oral and long-acting antipsychotics.

## INTRODUCTION

Schizophrenia is a chronic, complex and heterogenous mental disorder that presents with psychotic, disorganized, and negative symptoms. It has an estimated global prevalence of 0.28% (approximately 21 million individuals in 2016), peaking at 40 years old, affecting males and females almost equally, and with minimal variation across countries^1^. In the Philippines, schizophrenia has an estimated prevalence of 0.21% in 2017 – a figure approximately 25% higher than estimated prevalence in 1990 – with peak at age 35 to 39 years, and equal distribution between the sexes [2]. Burden attributable to schizophrenia is mainly from disability associated with the condition, which accounted for 1.7% of global years lived with disability (YLD) in 2016, and 1.42% of total YLD in the Philippines in 2017^1,2^. However, it has been previously noted that individuals with schizophrenia have about three times higher likelihood of dying when compared with the general population, usually from suicide or comorbid somatic conditions^3^.

Antipsychotic medications remain the mainstay for treatment for schizophrenia, which is usually offered in conjunction with psychosocial and family interventions and provided through a service delivery system that considers individual patient needs and the local context in which such treatment is provided^4^. Most clinical practice guidelines recommend the use of second-generation antipsychotic medications for first-episode schizophrenia^5^. The use of oral antipsychotics, however, has been found in both quantitative and qualitative studies to be associated with nonadherence in about 50% of cases, primarily relating to regimen, adverse effects, and cost of medications^6–8^. Nonadherence was found to have adverse consequences not only to patients (i.e., rehospitalization) but also to healthcare systems (i.e., increased cost)^7,8^.

Long-acting injectable (LAI) antipsychotics have been introduced in 1996 to address concerns encountered with oral antipsychotic medications^9–12^ and has since been incorporated in clinical practice guidelines for the treatment of schizophrenia^5,13,14^. This primarily relates to the demonstrated efficacy, safety and real-world effectiveness of LAIs when compared with oral antipsychotics, which was in turn attributed to, among others, better medication compliance^15^.

In the Philippines, however, no evidence has been published regarding the use of LAIs in an actual clinical setting. Given the unique context of the local health system, it is imperative to complement clinical trial data with real-world evidence on the use of LAIs. The introduction of paliperidone palmitate in a specialty hospital in 2018 provides an opportune moment to carry out an assessment of the medication’s impact not only to individual patients but to the healthcare institution as well.

Thus, this study was conceptualized to describe the clinical outcomes related to the introduction of paliperidone palmitate in a specialty hospital in the Philippines. Specifically, we set out to (a) describe the socio-demographic and clinical profile of patients given paliperidone palmitate in the specialty hospital; and (b) determine the clinical outcome of patients given paliperidone palmitate.

## METHODS

### Study design

We conducted a cross-sectional study among patients seen at a specialty hospital established in 1955 and currently under the Department of National Defense that offers medical care and treatment of the veterans and their dependents. The hospital, located in Metro Manila, provided paliperidone palmitate for free among eligible patients seen at the Department of Psychiatry since early 2018.

### Study participants

Participants of interest to this study were patients (a) age 18 to 65 years, (b) with a diagnosis of schizophrenia, (c) seen at the psychiatry service of the specialty hospital, and (d) who were initiated on paliperidone palmitate from March 2018 to June 2019. As part of baseline data were retrieved from medical records, patients with incomplete data specifying (a) primary diagnosis, (b) prior treatment used, and (c) outcome of treatment with paliperidone palmitate were excluded from the study. There are less than 100 patients given paliperidone palmitate in the specialty hospital, hence, all eligible patients (currently estimated to be around 60) were included. Using the sample size formula for estimating the population proportion, a minimum sample size of 38 patients diagnosed with schizophrenia who were given paliperidone palmitate is required to estimate the proportion of patients who experienced remission given the following information: 1) 50% of the patients are expected to have experienced remission; 2) margin of error set at 10%; 3) confidence level set at 95%; 4) design effect set at 1.0; and 5) the population size of patients given paliperidone palmitate in the specialty hospital is estimated at 60. The Open Epi website was used in computing the sample size. We originally planned for simple random sampling of prospective study participants. However, due to the pandemic situation, we recruited patients using convenience sampling (i.e., we invited those who presented at the out-patient department for their regular consultation/check-up).

### Data collection

Similar to the studies done previously^16,17^, we reviewed and abstracted baseline patient data from the medical record of eligible patients (i.e., last/latest recorded data prior to initiation of treatment with paliperidone palmitate) for the following variables: age, sex, place of residence, marital status, duration of illness, medical history, index drug, and date of initiation of treatment. Outcome of treatment was collected through a one-time objective assessment of the patient by a third-party psychiatrist (i.e., not involved in the care of the patient, hereinafter referred to as the “Sub-Investigator”) using the Structured Clinical Interview for Symptoms of Remission (SCI-SR), a tool that measures remission based on scores (1=absent, 8=present; a score of <3 in all items maintained over a six-month period indicate remission) on eight items of the Positive and Negative Syndrome Scale^18^. Assessment using the SCI-SR is estimated to take around 15 minutes or less.

### Data analysis

Data were encoded in Microsoft Excel and analyzed using Stata/MP 14.2 for Mac. Continuous variables (e.g. age, length of stay, etc.) were described using mean and standard deviation, while categorical variables (e.g. sex, type of past medications, clinical outcomes, etc.) were summarized using frequency and percentage distribution tables. To answer the second specific objective of this study, point and 95% confidence interval estimates of the proportion of patients who experienced schizophrenia remission was computed.

### Ethical considerations

This research was conducted in accordance with the principles laid out in the *National Ethical Guidelines for Health and Health-related Research 2017*, and in compliance with the provisions of the Data Privacy Act of 2012 as well as other relevant statues and regulations promulgated by duly constituted authorities. This research protocol was reviewed and approved by the Institutional Review Board of the Veterans Memorial Medical Center prior to implementation (VMMC-2020-015).

## RESULTS

A total of 30 patients were recruited for the study from August 2020 and June 2021. This represents 80% of the minimum sample size computed for the study – a decent number that allows us to draw inferences from the data – and 50% of the estimated population of individuals treated with paliperidone palmitate who fit our eligibility criteria. A principal reason for not being able to attain full recruitment was that the data collection period coincided with the imposition of community quarantine measures as part of the government’s pandemic control policy. This hindered patients, especially those living outside of Metro Manila, from accessing the hospital’s outpatient department. Further, some patients who sought consult at the institution, while treated with paliperidone palmitate, did not fully meet our eligibility criteria.

Profile of patients given paliperidone palmitate. Of the thirty patients who participated in the study, the majority were males (80%), residents of the National Capital Region (50%), and single (20%) (Table 1).

**Table 1.** Sociodemographic profile of participants

| Characteristics | n (%) |
| --- | --- |
| Sex |  |
| Female | 5 (16.67) |
| Male | 24 (80.00) |
| *Not indicated* | 1 (3.33) |
| Residence |  |
| National Capital Region |  |
| Quezon City | 5 (16.67) |
| Makati City | 3 (10.00) |
| Pasay City | 2 (6.67) |
| Marikina City | 1 (3.33) |
| Muntinlupa City | 1 (3.33) |
| Pasig City | 1 (3.33) |
| Taguig City | 1 (3.33) |
| Valenzuela City | 1 (3.33) |
| Region 3 |  |
| Bulacan | 2 (6.67) |
| Nueva Ecija | 1 (3.33) |
| Region 4 |  |
| Antipolo City | 5 (16.67) |
| Cavite | 2 (6.67) |
| Laguna | 2 (6.67) |
| Rizal | 2 (6.67) |
| Batangas | 1 (3.33) |
| Marital status |  |
| Single | 20 (66.67) |
| Married | 4 (13.33) |
| Separated | 5 (16.67) |
| *Not indicated* | 1 (3.33) |

As shown in Table 2, the most common behavioral risk factor reported was smoking (20%). In terms of psychiatric comorbidities, one participant was each reported to have anxiety and depressive symptoms. On the other hand, the most common medical comorbidities reported were hypertension (30%) and diabetes mellitus (16.67%).

**Table 2.** Risk factors and comorbidities

| Characteristics | n (%) |
| --- | --- |
| Behavioral / Metabolic risk factors |  |
| Smoking | 6 (20.00) |
| Obesity | 2 (6.67) |
| Alcohol use | 1 (3.33) |
| Psychiatric comorbidities |  |
| Anxiety | 1 (3.33) |
| Depressive symptoms | 1 (3.33) |
| Medical comorbidities |  |
| Hypertension | 9 (30.00) |
| Diabetes mellitus | 5 (16.67) |
| COPD | 3 (10.00) |
| Thyroid disorder | 1 (3.33) |

NOTE: Multiple responses allowed for all items

All the participants were treated with paliperidone for schizophrenia. The median duration from schizophrenia diagnosis to initiation of paliperidone treatment was 19.50 years (IQR: 16.60 – 33.50), while the median age of participants at the initiation of paliperidone treatment was 52 years (IQR: 46 – 55) (Table 3). Prior to the initiation of paliperidone treatment, the most common antipsychotic drugs taken by participants were clozapine (50%), haloperidol (26.67%), and olanzapine (16.67%). In eight patients (22.67%), other antipsychotic drugs were discontinued following initiation of paliperidone treatment; in the remaining 22 participants (73.33%), paliperidone was taken concurrently with other antipsychotic drugs.

**Table 3.** Participant characteristics related to paliperidone treatment

| Characteristics | n (%) |
| --- | --- |
| Duration from schizophrenia diagnosis to initiation of paliperidone treatment (years) |  |
| Median (IQR) | 19.50 (16.50 – 33.50) |
| Mean ± SD | 22.43 ± 10.95 |
| Age at start of paliperidone treatment (years) |  |
| Median (IQR) | 52 (46 – 55) |
| Mean + SD | 49.27 ± 9.78 |
| Psychiatric drugs taken prior to initiation of paliperidone treatment* |  |
| Clozapine | 15 (50.00) |
| Haloperidol | 8 (26.67) |
| Olanzapine | 5 (16.67) |
| Risperidone | 4 (13.33) |
| Quetiapine | 3 (10.00) |
| Flupentixol | 3 (10.00) |
| Valproate | 1 (3.33) |
| Sertraline | 1 (3.33) |
| Current drug regimen |  |
| Paliperidone only | 8 (26.67) |
| Paliperidone + other psychiatric drugs | 22 (73.33) |

NOTE: \*Multiple responses allowed for this item

Clinical outcome of patients given paliperidone palmitate. Response to paliperidone treatment was defined in terms of remission, which was measured using the SCI-SR. SCI-SR is a tool that rates each of eight symptoms using a seven-point Likert scale (1 = absent to 7 = severe). A participant was classified to be in remission if all the individual scores is 3 or lower (i.e., absent to minimal), at least 6 months from the initiation of treatment.

The median duration from the initiation of paliperidone treatment to follow-up assessment was 27.20 months (IQR: 24.73 – 30.50), with all participants having at least 6 months of treatment (Table 4). At follow-up assessment, all participants were classified to be in remission.

**Table 4.**
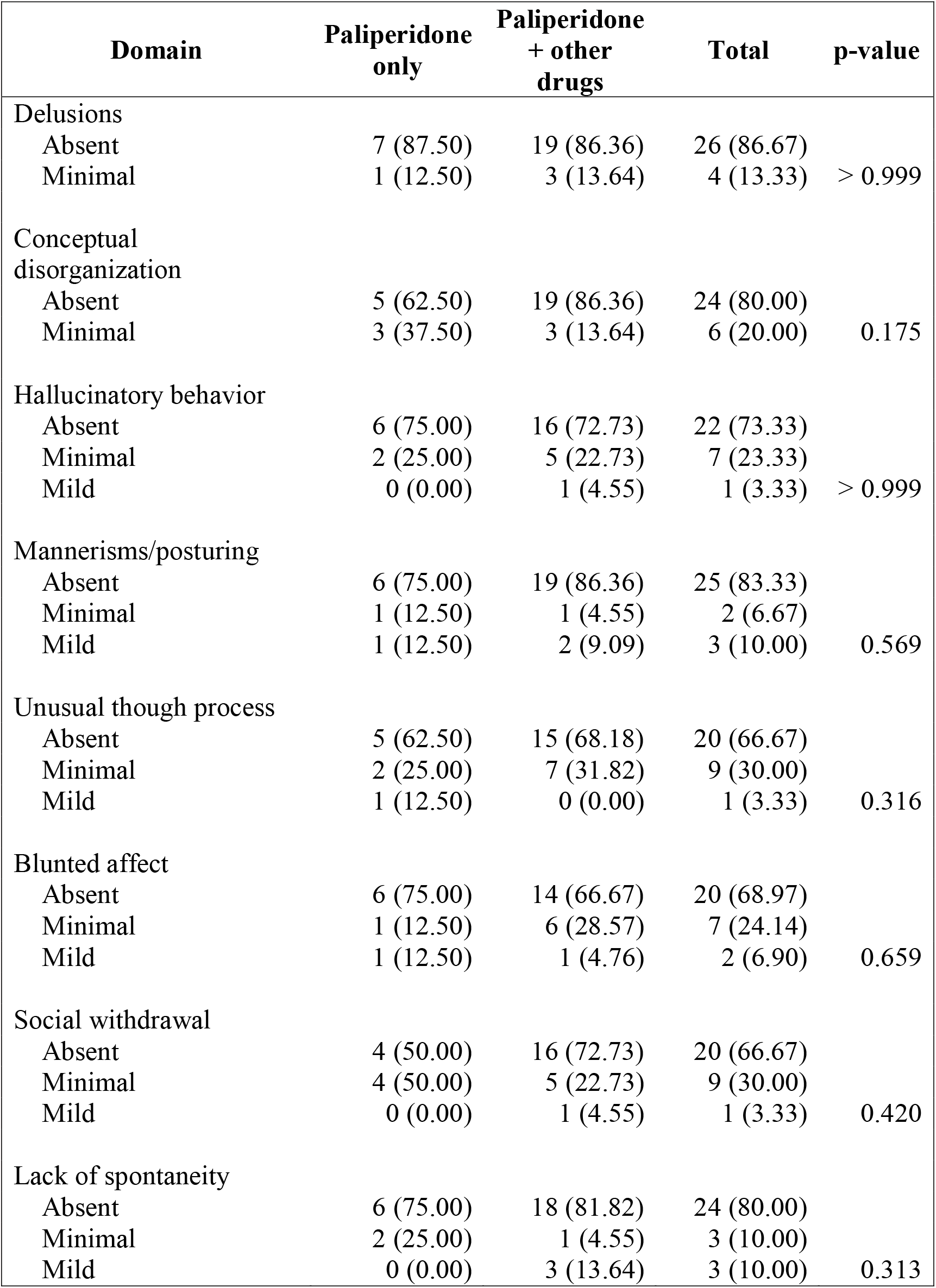
Comparison of response to paliperidone treatment, by treatment regimen and overall

Since participants had differing paliperidone treatment regimens (i.e., paliperidone only versus paliperidone + other drugs), we compared the distribution of scores for each symptom included in the SCI-SR between participants defined by treatment regimen. Our analyses indicate that there is no significant difference in SCI-SR scores between those who took paliperidone only versus those who took paliperidone concurrently with other drugs.

Two participants were reported to have subcutaneous nodules at the injection site, and this adverse event was assessed by the attending psychiatrist as mild in severity. In both participants, the subcutaneous nodules were managed with warm compress on the affected site; however, there was reportedly no immediate resolution of adverse event.

## DISCUSSION

This study was conducted to describe the clinical outcomes related to the introduction of paliperidone palmitate in a specialty hospital in the Philippines. All 30 patients with schizophrenia who were given paliperidone palmitate (alone or in combination with other oral antipsychotic drugs) were found to be in remission at the time of the study.

Our results contribute to evidence on the real-world effectiveness of paliperidone palmitate for the treatment of schizophrenia reported elsewhere^15^. However, there is a degree of uncertainty with this finding since, while all patients were in remission at the time of assessment, only a handful were given paliperidone palmitate alone, and we found no significant difference in terms of reported outcomes among those given paliperidone palmitate alone compared to those on combination treatment. This is consistent with earlier systematic reviews^19,20^ which showed no significant difference in symptoms and relapse among patients treated with either long-acting injectable antipsychotics and oral antipsychotics.

One interpretation of this finding is that paliperidone palmitate alone may be effective in achieving remission among patients with schizophrenia, thus averting the need for polypharmacy and reducing treatment costs (estimated to be around Php 470,000, or USD 9,700, per annum if the specialty hospital will shift all patients from oral antipsychotics to paliperidone palmitate).

This positive outcome was also cited by patients and health workers in the institution as an important factor in encouraging patients and health workers alike to continue the use of paliperidone palmitate.

An important limitation of this study is that it is challenging to conclusively state that paliperidone palmitate led to remission of patients included in this study. As previously mentioned, there was no significant difference in remission outcomes for patients who were given paliperidone palmitate alone compared to those on combination treatment. Further, baseline assessment, while available in the medical records, did not use SCI-SR, precluding comparability of patient status before and after treatment with paliperidone palmitate.

## CONCLUSION

In this study among patients with schizophrenia seen in a specialty hospital in the Philippines, we found evidence that clinical outcomes with paliperidone palmitate were comparable to those given a combination of oral and long-acting antipsychotics.

## Data Availability

All data produced in the present study are available upon reasonable request to the authors.

